# Reliability of Spike Gene Target Failure for ascertaining SARS-CoV-2 lineage B.1.1.7 prevalence in a hospital setting

**DOI:** 10.1101/2021.04.12.21255084

**Authors:** José Afonso Guerra-Assunção, Paul A. Randell, Florencia A. T. Boshier, Michael A. Crone, Juanita Pang, Tabitha Mahungu, Paul S. Freemont, Judith Breuer

## Abstract

The appearance of the SARS-CoV-2 lineage B.1.1.7 in the UK in late 2020, associated with faster transmission, sparked the need to find effective ways to monitor its spread. The set of mutations that characterise this lineage include a deletion in position 69 and 70 of the spike protein, which is known to be associated with Spike Gene Target Failure (SGTF) in a commonly used three gene diagnostic qPCR assay. The lower cost and faster turnaround times compared to whole genome sequencing make the use of qPCR for monitoring of the variant spread an attractive proposition. However, there are several potential issues with this approach. Here we use 826 SARS-CoV-2 samples collected in a hospital setting as part of the Hospital Onset COVID Infection (HOCI) study where qPCR was used for viral detection, followed by whole genome sequencing (WGS), to identify the factors to consider when using SGTF to infer lineage B.1.1.7 prevalence in a hospital setting, with potential implications for locations where this variant has recently been introduced.

## Introduction

The emergence of the UK Variant of Concern (VOC) 202012/01, also known as lineage B.1.1.7, in South East England during the latter part of 2020 has been associated with rising rates of transmission [1] and potentially with increased disease severity [2].

Increased prevalence of this variant is correlated with an increase in the number of qPCR-based community diagnostic tests that fail to detect spike (S) gene amplicons [3]. So called spike gene target failure (SGTF) has been shown to be due to a deletion of amino acids 69 and 70 in the spike protein leading to failure of probes to bind to the S gene amplicon in one commercial qPCR assay. Detection of other amplicon targets, including the nucleocapsid (N) and ORF1ab genes is unaffected. In community studies SGTF shows good correlation of SGTF with lineage B.1.1.7 after mid-November, but is less accurate before that date [3].

To determine how well SGTF corresponded to VOC in patients hospitalised over the same period we made use of data collected as part of the COVID-19 UK Hospital Onset COVID Infection (COG-UK-HOCI). Only one of 15 hospitals in this trial is using an assay that involves S gene target detection in inpatients while another uses it for healthcare workers only. In this dataset we present data on 826 samples collected between weeks 41 and 53 of 2020. This includes 535 hospital patients collected from week 41, and 291 healthcare workers collected from week 49.

## Results

An initial analysis based on unreported S gene results was undertaken. The numbers of samples with and without SGTF from two laboratories are shown in Table 1 and the increasing proportion of SGTF, over time from weeks 45-53 is shown in Figure 1. Among samples with SGTF the proportion due to lineage B.1.1.7, as determined by sequencing, started at 0% in week 43, rising to 20% by week 46 and to nearly 100% by week 53 (Figure S1 and S2). The majority (37/49) of SGTF that were not lineage B.1.1.7 were lineage B.1.258, which is known to carry the same 69/70 two amino acid deletion. SGTF was also observed in 12 samples of other lineages that do not have 69/70 S gene deletion (Table 2).

**Table 1.** Number of samples from each laboratory with a breakdown by lineage status as identified by pangolin.

|  | VOC202012/01 | Non VOC |
| --- | --- | --- |
| IHT | 143 | 392 |
| RFH | 218 | 73 |

**Table 2.** Breakdown of samples that demonstrate SGTF in this dataset by pangolin lineage and presence of deletion of spike amino acids 69 and 70.

| Lineage | Spike 69/70del | Count |
| --- | --- | --- |
| B.1.1.7 | Y | 337 |
| B.1.258 | Y | 37 |
| B.1.177 | N | 7 |
| B.1.5 | Y | 2 |
| B.1.102 | Y | 1 |
| B.1.19 | Y | 1 |
| B.1.5 | N | 1 |

**Figure 1.**
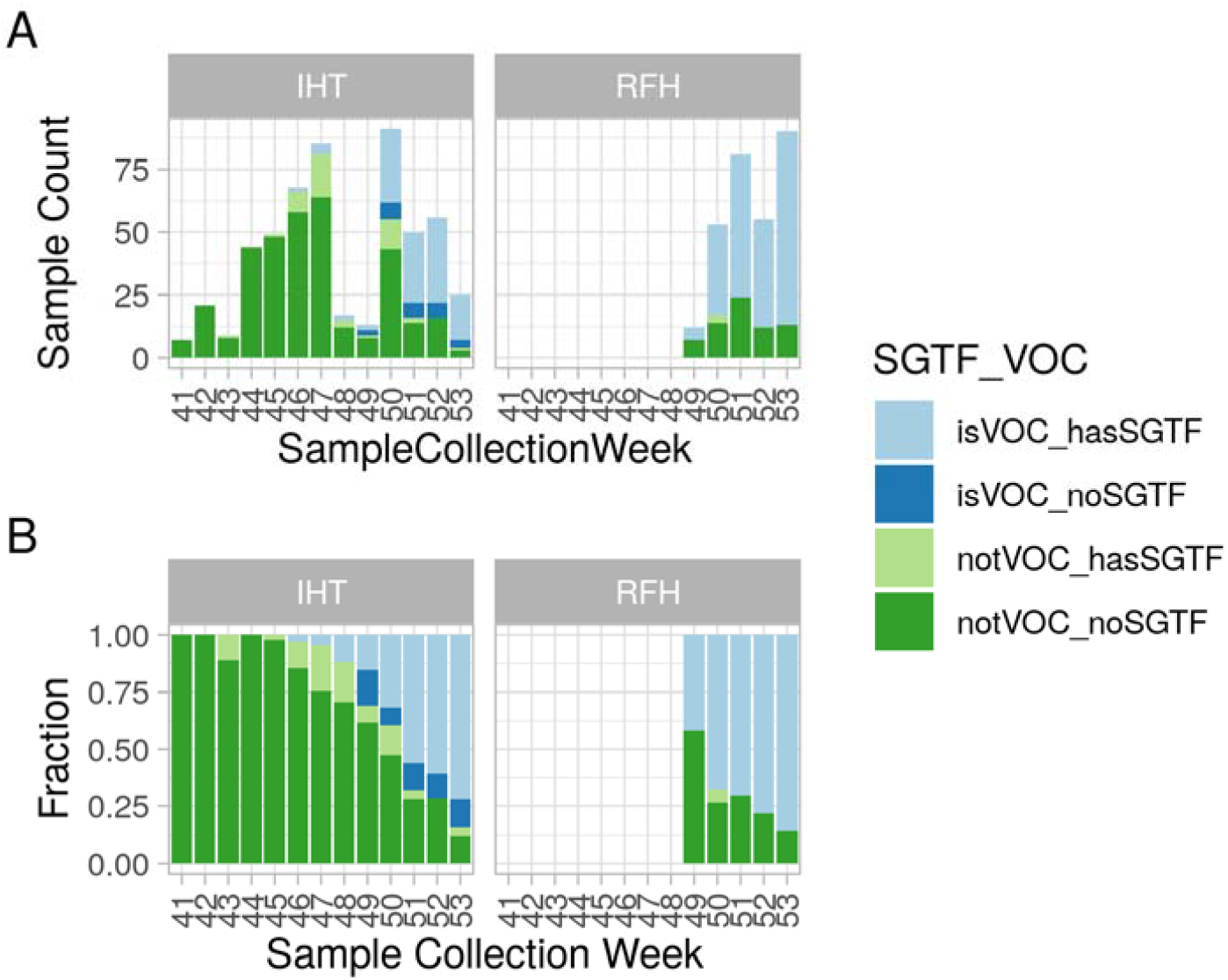
Number of samples sequenced in each week (A) for each of the hospital trusts in this study. Samples that are lineage B.1.1.7 are shown in blue, while samples from other lineages are shown in green. Samples that demonstrate SGTF are shown in a lighter colour than those that do not demonstrate SGTF. Panel B shows the fraction of samples in each category.

Surprisingly, an apparent signal from the S-gene (based on the autogenerated CT values) was present in 17% (24 out of 143) of samples later confirmed as lineage B.1.1.7s by sequencing, all of which had deletions at amino acids 69/70 (Figure 1 and S3). Closer scrutiny of the amplification curves revealed that in these samples the S-gene curves were not consistent with genuine amplification curves as demonstrated in Figure 2. Although the curves crossed the auto-threshold, these would be reported as S-gene not detected if individual targets were being reported. Without manual curation these samples were falsely identified as not lineage B.1.1.7, resulting in underestimation of the true lineage B.1.1.7 prevalence. To further investigate this phenomenon, we compared the qPCR CT values for each of the three genes for lineage B.1.1.7 with apparent SGTF with those samples demonstrating the anticipated SGTF (Figure 3). Samples with the spurious signal for the S gene had significantly lower N and ORF1ab CT values (compared to samples with SGTF) and within this group the S gene CT values were always significantly higher than the CT values for the other genes (p<0.001).

**Figure 2.**
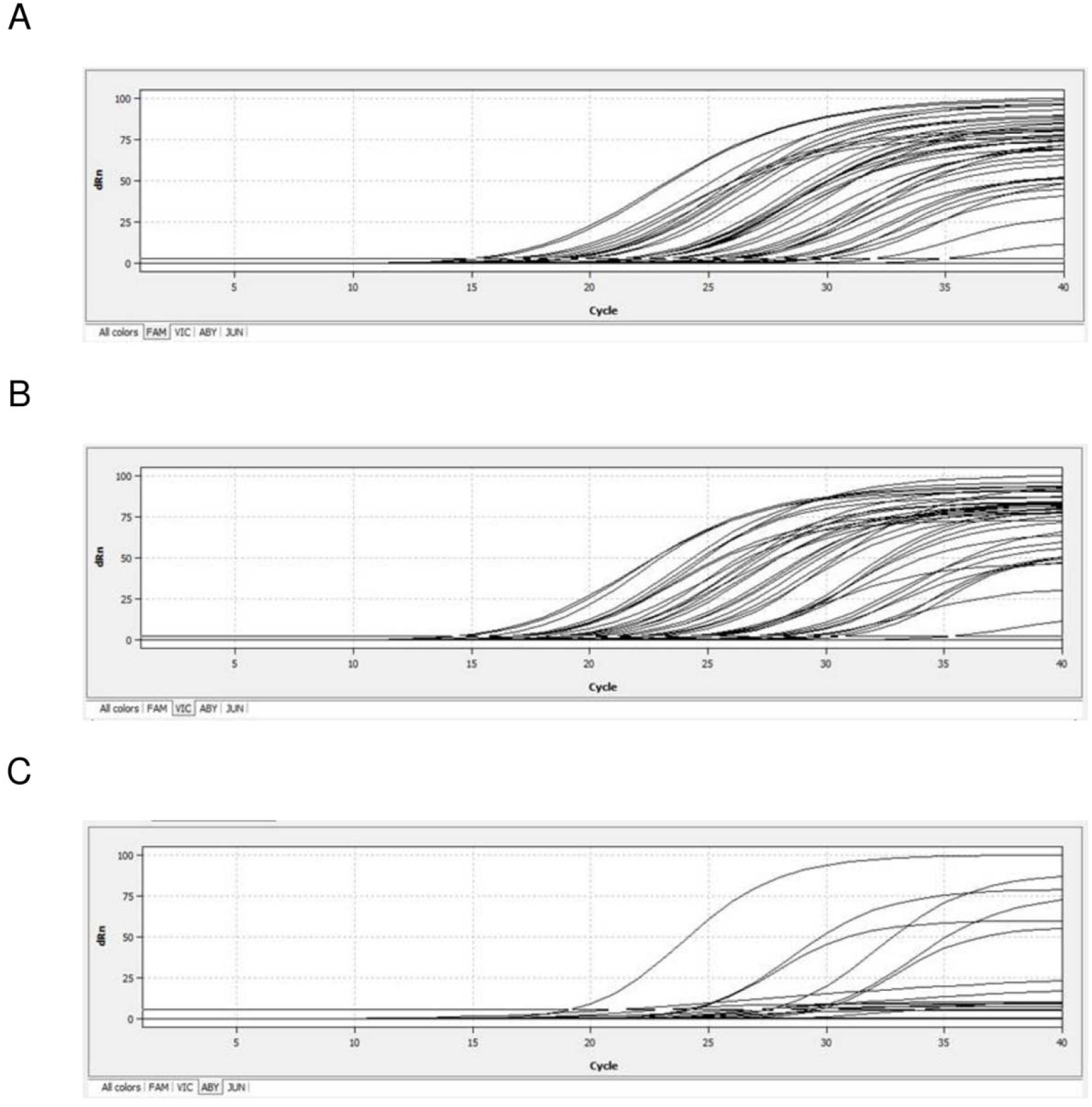
Amplification curves obtained for a subset of samples in this study. Each panel corresponds to a different gene: A) ORF1 ab, B) N gene, C) S gene, where some flattened amplification curves that cross the automatic threshold for detection can be seen.

**Figure 3.**
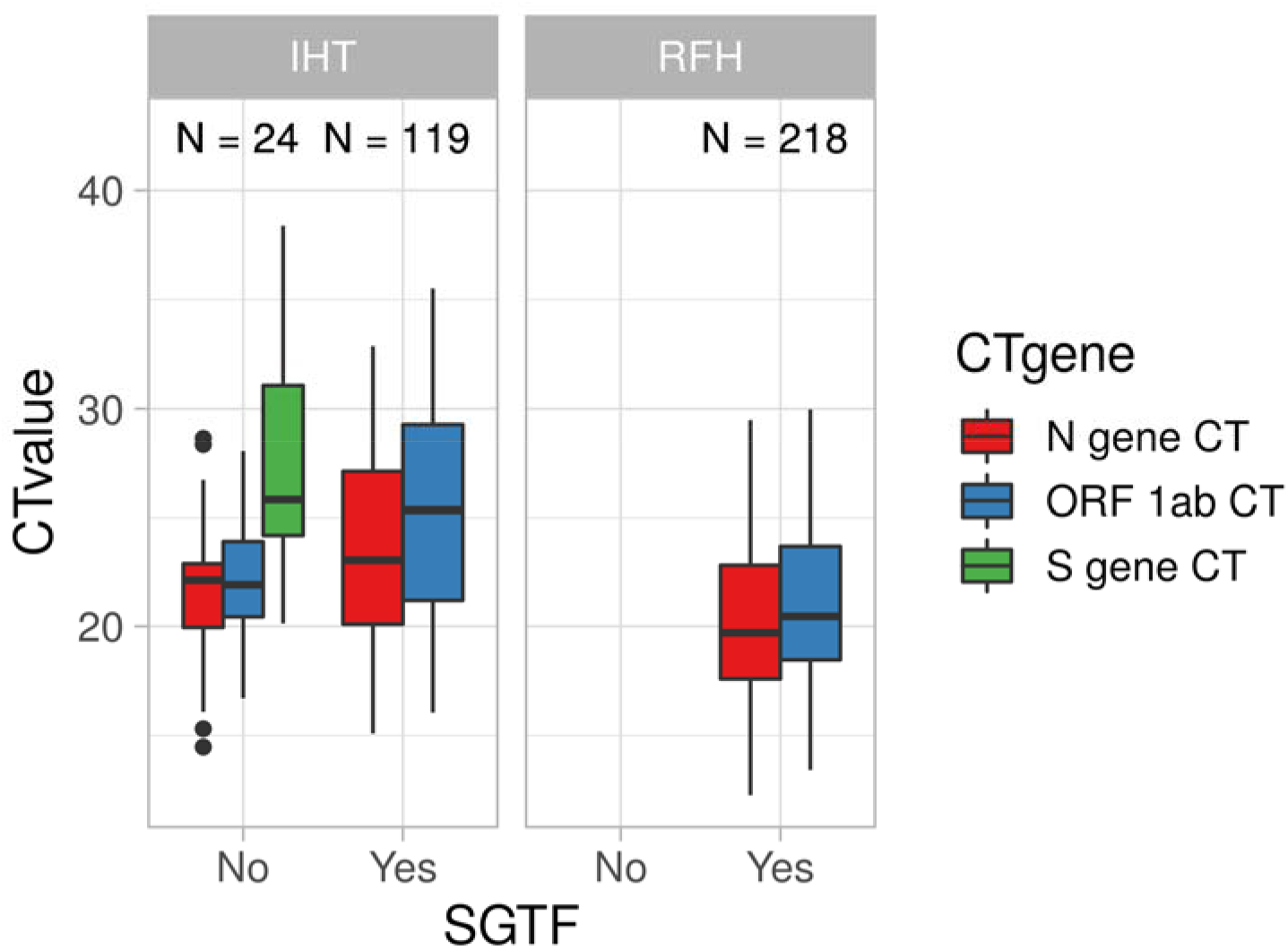
Comparison between the CT values of lineage B.1.1.7 samples that demonstrate SGTF in comparison to those that do not. Each gene in the assay is represented by a different boxplot (N in red, ORF1ab in blue, S in green). The number of samples in this dataset for each category is shown above each boxplot pair. The RFH health trust has no lineage B.1.1.7 samples that do not show SGTF. In the IHT samples, it is clear that the CT value of the S gene is significantly higher than the other two genes in the assay (p < 0.001).

To investigate whether the viral load was a factor in this phenomenon a subset of samples including lineage B.1.1.7 samples that had previously not shown SGTF, samples that had shown SGTF as expected and samples from other lineages without the relevant deletion were retested neat and when serially diluted. The spurious S gene signal was not reliably reproducible and did not appear to be related to the viral load (data not shown).

## Discussion

SGTF has been used to identify lineage B.1.1.7 in the community and forms the basis for many studies looking at its transmission and severity [1,4–6]. Our data suggest that both false positive and false negatives can occur which may skew the positive predictive value of SGTF. In this study, lineage B.1.258, which also carries the spike deletion 69/70, was prevalent locally and we observed false positive lineage B.1.1.7 calls in 9.6% of cases. The incidence of false positives decreased as the more transmissible B.1.1.7 outcompeted B.1.258. By week 51 more than 90% of SGTF was B.1.1.7 (Figure S2).

We also found that there is a risk that spurious S gene results may be observed (as was the case with one of the assays used) and without careful review and consideration of the PCR results the prevalence of SGTF (and consequently lineage B.1.1.7) may be underestimated. This could be due to imperfect colour compensation on qPCR machines other than those made by the manufacturer of the assay, further complicated by the lack of controls for each single target of the multiplex assay to check for colour bleedthrough. The qPCR data from the manufacturer’s machine is also interpreted by separated software developed by the manufacturer and it is unclear how thresholding is performed and how the software handles any noise or curves that do not have an exponential shape.

This phenomenon is evidently dependent on the PCR machine and dyes used. This is reflected in the fact that the false negative results are restricted to just one of the two sites studied (one site used the manufacturer’s qPCR machine, the other a different machine with colour compensation performed with the manufacturer’s calibration plates).

While this phenomenon was only observed in one of the two set of samples analysed here, it has also been independently observed for lineage B.1.1.7 and SGTF in Portugal. [7]. Additionally, a locally developed assay that mimicked SGTF with a different dye did not show the same behaviour (data not shown).

Thus, in summary, SGTF is an important surrogate marker for the VOC 202012/01, and useful for large scale epidemiology studies. However, care needs to be taken where sample numbers are small, for example in care homes or hospitals for example where the intention is to link lineage B.1.1.7 to outcomes or in scenarios where lineage B.1.1.7 variants are in the minority. In these cases, the use of SGTF can be misleading and sequencing is recommended.

## Data Availability

Sequencing data was deposited when generated to the appropriate SARS-CoV-2 public repositories (COG-UK and GISAID) and is publicly available.

## Acknowledgements

Data for this study was collected as part of the COG-UK HOCI study. COG-UK HOCI is part of COG-UK. COG-UK is supported by funding from the Medical Research Council (MRC) part of UK Research & Innovation (UKRI), the National Institute of Health Research (NIHR) and Genome Research Limited, operating as the Wellcome Sanger Institute.

## Materials and Methods

### Sequencing

Whole genome sequences for SARS-CoV-2 were generated following a positive qPCR test using either Illumina or ONT nanopore technologies, according with COG-UK [8] processes and deposited in the appropriate repositories. Lineages were determined using Pangolin [9]. The deletion of Spike amino acids 69 and 70 was independently inspected after alignment to reference using minimap2 [10]. The data was aggregated, analysed and visualised with the R statistical framework.

### RFH qPCR method

Viral samples are inactivated using RNA Inactivation buffer, and RNA is extracted using an automated in-house method, based on a protocol previously described [11].

The extracted RNA undergoes a RT-qPCR assay and is quantified on a ABI QuantStudio5 system together with a positive control (TaqPath COVID-19 Control) and an extraction control.

Presence of viral targets is indicated by an S shape curve with CT < 37, together with concordant duplicates and the presence of the MS2 control trace at CT < 32

### IHT qPCR methods

A sample volume of 200 μl was used for RNA extraction using the Maxwell HT Viral TNA kit (Promega) with a custom extraction protocol [12] on the CyBio FeliX liquid handler (Analytik Jena) with an elution volume of 50 μl. Subsequent RT-qPCR was performed using the TaqPath™ COVID-19 CE- IVD RT-PCR Kit (ThermoFisher Scientific) according to the manufacturer’s instructions and thermocycled on a qTower3 (Analytik Jena). Colour compensation was performed using FAM, VIC, ABY and JUN dyes (ThermoFisher Scientific) according to the qTower3’s calibration instructions. When determining the CT values for the different targets the auto-threshold generated by the analyser was used. Samples were reported as SARS CoV-2 RNA detected when at least 2 targets were detected with typical exponential growth curves. Samples not meeting these criteria were reported as not detected if no targets were present or underwent confirmatory testing if there was only one target. As the overall result was reported (and not individual targets) the autogenerated CT values were not manually curated.

## Supplementary Figures

**Figure S1.**
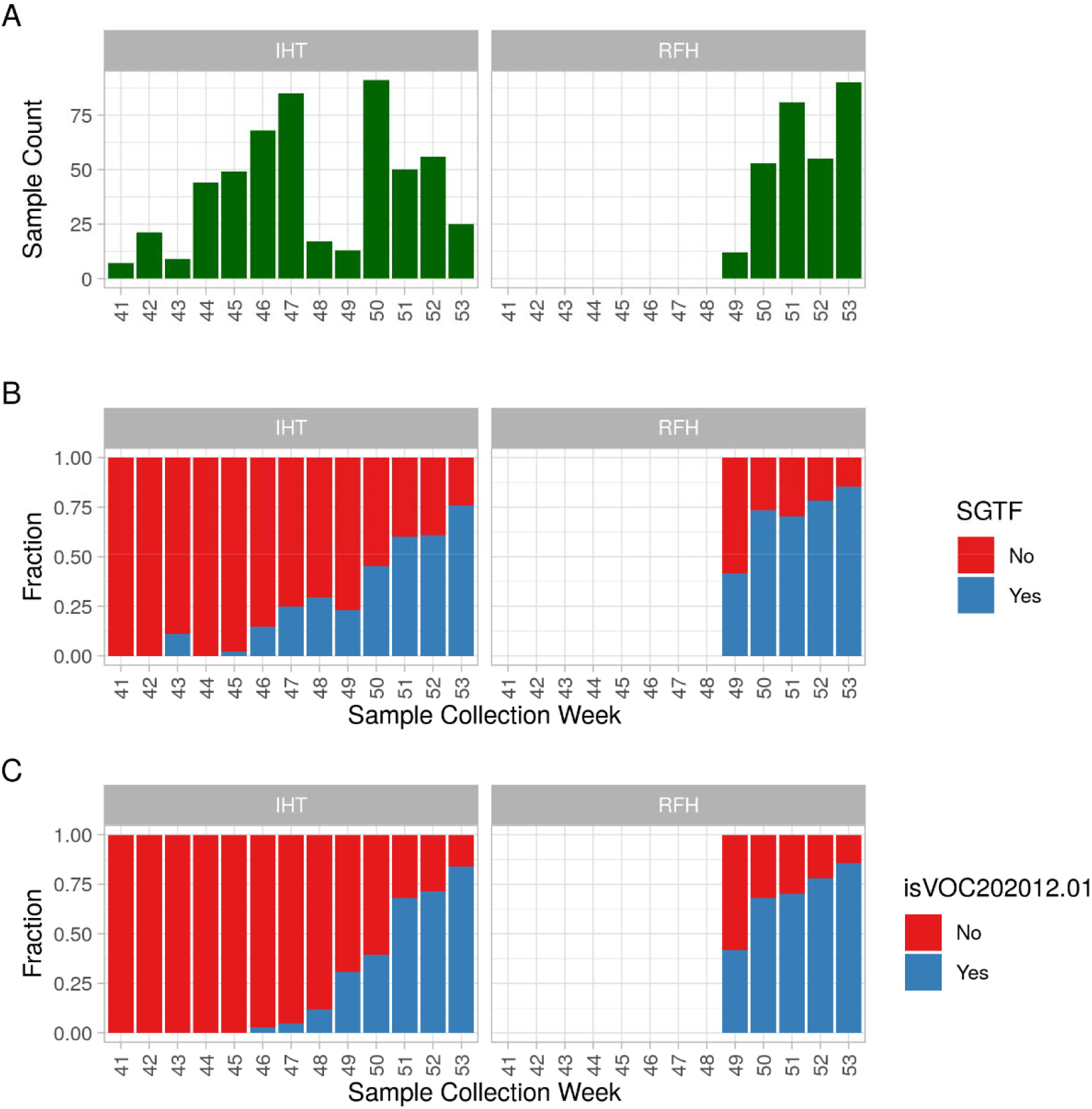
Total number of samples in this study stratified by week of sample collection and laboratory where they originated from. (A) shows the total number of samples for each week. (B) illustrates the fraction of samples that demonstrate SGTF (blue) or do not demonstrate SGTF (red) across time. (C) illustrates the fraction of samples that were classified by pangolin as lineage B.1.1.7 (blue) or other lineage (red) across time.

**Figure S2.**
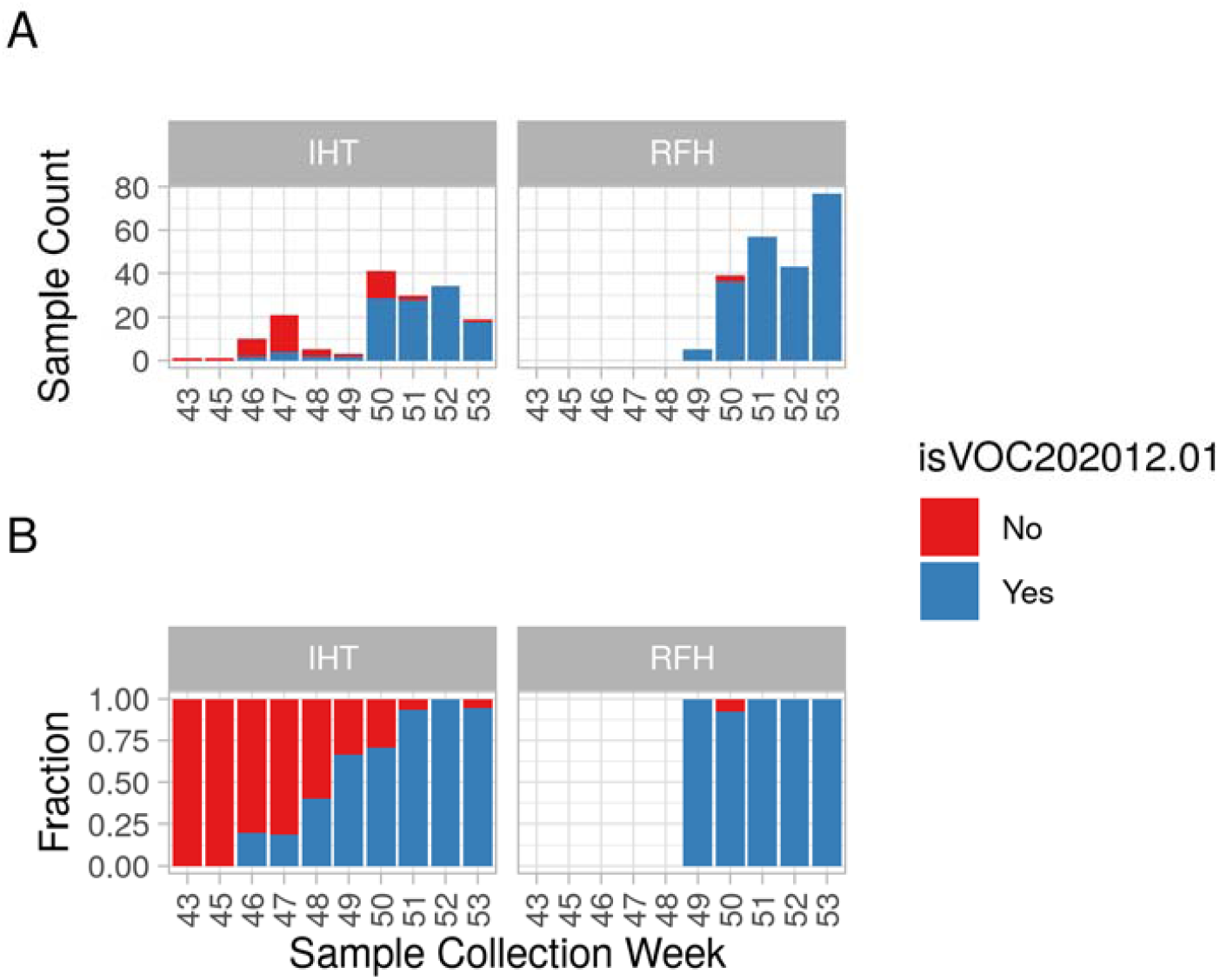
Number of SGTF instances by week (A) for each of the hospital trusts in this study, and fraction of SGTF samples per week that are lineage B.1.1.7 (B). SGTF events are separated between those samples that are not lineage B.1.1.7 (red) and those that are lineage B.1.1.7 (blue).

**Figure S3.**
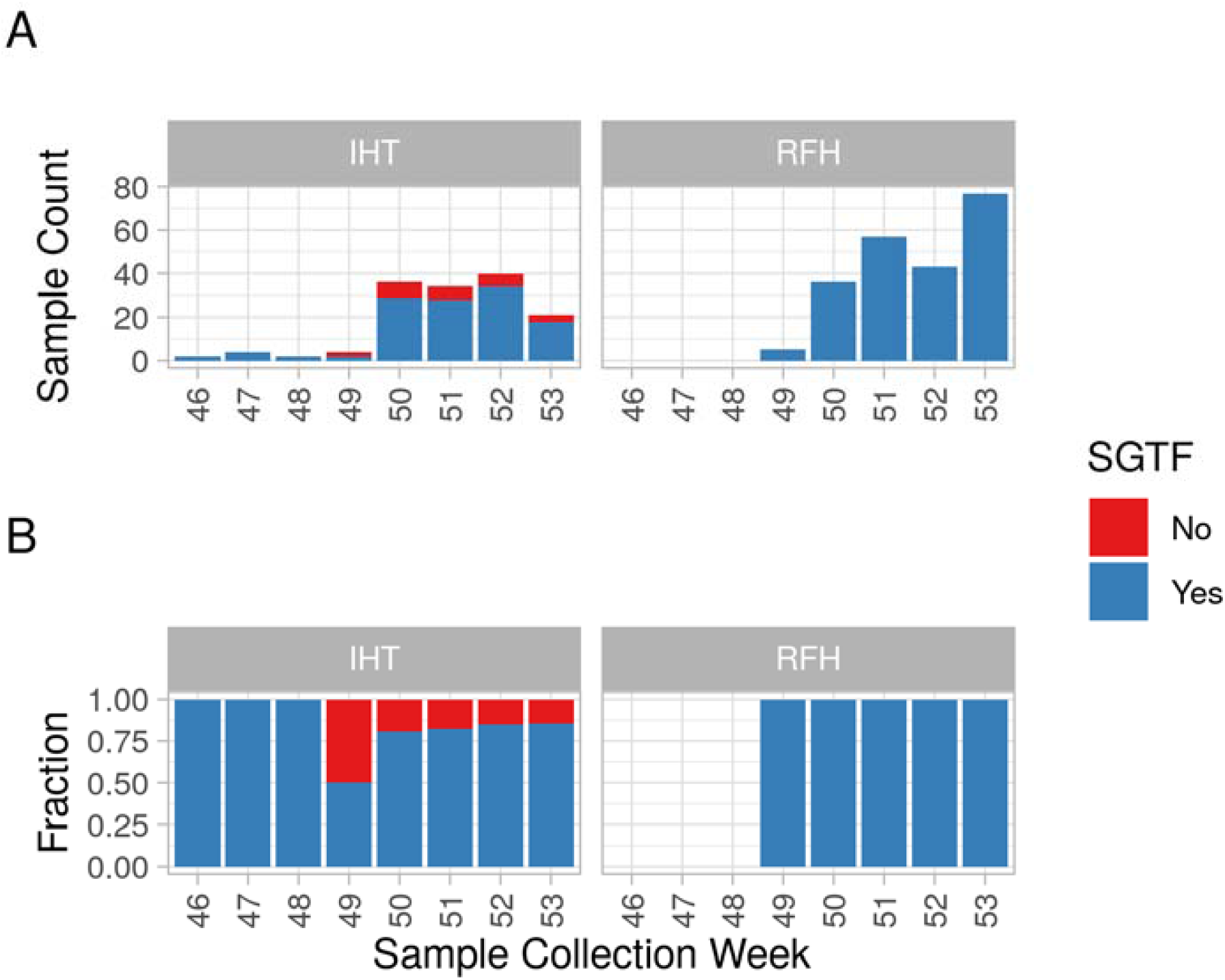
Number of lineage B.1.1.7 samples by week (A) for each of the hospital trusts in this study, and fraction of lineage B.1.1.7 samples per week that cause SGTF (B). Lineage B.1.1.7 events are separated between those samples that did not cause SGTF (red) and those that have undetectable S gene (blue).

